# Predicting the Efficacy of COVID-19 Convalescent Plasma Donor Units with the Lumit Dx anti-Receptor Binding Domain Assay

**DOI:** 10.1101/2021.03.08.21253135

**Authors:** Sanath Kumar Janaka, Natasha M Clark, David T Evans, Joseph P Connor

**Author notes:** Corresponding Author Joseph P. Connor, MD, 3147 MFCB 1685 Highland Ave., Madison, WI 53705. Equal contribution (Sanath Kumar Janaka, Natasha M Clark). Reagents for Lumit Dx assays were provided by Promega for this study. The authors have no other conflicts to report.

## Abstract

**Background:** The novel coronavirus SARS-CoV2 that causes COVID-19 has resulted in the death of more than 2.5 million people, but no cure exists. Although passive immunization with COVID-19 convalescent plasma (CCP) provides a safe and viable therapeutic option, the selection of optimal units for therapy in a timely fashion remains a barrier.

**Study design and methods:** Since virus neutralization is a necessary characteristic of plasma that can benefit recipients, the neutralizing titers of plasma samples were measured using a retroviral-pseudotype assay. Binding antibody titers to the spike (S) protein were also determined by a clinically available serological assay (Ortho-Vitros total IG), and an in-house ELISA. The results of these assays were compared to a measurement of antibodies directed to the receptor binding domain (RBD) of the SARS-CoV2 S protein (Promega Lumit Dx).

**Results:** All measures of antibodies were highly variable, but correlated, to different degrees, with each other. However, the anti-RBD antibodies correlated with viral neutralizing titers to a greater extent than the other antibody assays.

**Discussion:** Our observations support the use of an anti-RBD assay such as the Lumit Dx assay, as an optimal predictor of the neutralization capability of CCP.

## Introduction

Since its emergence in late 2019 the novel coronavirus SARS-CoV2 has crippled healthcare systems and taken the lives of at least 2.5M. With no consistently effective therapeutic interventions, the use of COVID-19 Convalescent Plasma (CCP) remains a viable treatment option (1-3). Passive immunization relies on neutralizing antibodies that can prevent infection of respiratory epithelial cells by the virus. Although verification of virus neutralization is the gold standard for determining the therapeutic value of any single unit of CCP (4), this is not practical for the scale of the pandemic. Alternatively, antibody measurements by commercial serological assay systems currently available correlate modestly with the neutralization activity of CCP (5-7). In general, and supported by data from our lab (8), the presence of anti-spike (S) protein antibodies is more predictive of virus neutralization than antibodies directed to other viral antigens. Among these, antibodies targeting the S protein receptor binding domain (RBD) in theory offer the best chance of viral neutralization by preventing interaction of the S protein with the ACE2 receptor on epithelial cells (9). Here, a novel single step immunoassay, Lumit Dx SARS-CoV-2 (Promega), was used to measure RBD specific immunoglobulins in 111 convalescent plasma donors. Neutralizing titers for these samples were also determined using a retroviral pseudotype assay (10). These antibody measures were then compared with total anti-S Ig measured with a commercial serological assay and anti-S IgG measured with in-house ELISAs.

## Materials and Methods

### ELISA

A modified ELISA, based on protocols published by Robbiani et al. and Routhu et al. (5, 21), were used to evaluate antibody binding to SARS-CoV-2 spike protein extracellular domain. 96-well plates were coated with 1µg/mL SARS-CoV-2 spike protein (S1+S2 ECD, Sino Biologicals) in PBS and stored at 4°C overnight. Plates were washed twice with wash buffer (PBS with 0.05% Tween-20) and incubated with blocking buffer (PBS, 5% nonfat milk, 1% FBS, and 0.05% Tween-20). Plates were washed and serum/plasma samples were added at a 1:200 starting dilution followed by 7 threefold serial dilutions. Rhesus Anti-SARS CoV Spike monoclonal antibody (NHP Reagent Resource), anti-Dengue monoclonal antibody, and 6 negative control plasma samples were added to each plate for validation. After 1h at 37°C, plates were washed and incubated with anti-human IgG secondary antibody conjugated to horseradish peroxidase (HRP) (Jackson Immunoresearch) in blocking buffer (1:5000 dilution) at 37°C for 1 h. TMB substrate was added (ThermoFisher), and absorbance read at 405 nm with a plate reader (Victor X4, Perkin Elmer). To evaluate IgA and IgM binding levels, the same plates were washed and incubated with HRP inhibitor (0.02% sodium azide in blocking buffer) for 30min. Complete loss of absorbance at 405 nm with TMB under these conditions was confirmed during assay development. Plates were washed before incubating with anti-human IgA or IgM conjugated to HRP (1:5000 dilution) (Jackson Immunoresearch) and absorbance data collected. Area under the curve (AUC) was calculated (Graphpad Prism) as a measure of the anti-S antibody titers.

### Lumit Dx Immunoassay

The Promega Lumit Dx SARS-CoV-2 Immunoassay was used according to the manufacturer’s protocol for detection of antibody binding to the RBD of SARS-CoV-2 spike protein. The assay utilizes luminescence to determine antibody binding to SARS-CoV-2 RBD and was measured using a Victor X4 plate reader (Perkin Elmer).

### Neutralization Assay

The neutralization assay was described previously (8, 10). 293T-ACE2 cells were seeded in 96 well plates. The following day, SARS-CoV-2 convalescent serum or plasma was diluted in D10 media (DMEM plus 10% FBS, 100 U/ml penicillin, 100 µg/ml streptomycin, 0.25 µg/ml amphotericin B, and 2mM L-glutamine) and mixed with retroviral particles that package a firefly luciferase reporter and pseudotyped with the SARS-CoV-2 spike protein. 1h later, the mixture was incubated with 293T-ACE2 cells for 48h. The cell lysates were then measured for luciferase activity with the Perkin Elmer Britelite system and the Victor X4 plate reader. Relative infectivity was determined by normalizing serum or plasma treated cells to those of no serum or plasma controls after subtracting signal from uninfected cells. The dilution at which the sample reduced infectivity to half the maximum was determined and reported as IC50 values.

## Results

Serum or plasma samples from 111 CCP donors provided by the American Red Cross (ARC) were assayed for anti-SARS-CoV-2 S protein antibodies by the assays described. Regardless of the assay, the measures of anti-spike antibodies were highly variable. Neutralization across the samples also varied with a wide range of half-maximal neutralization titers, IC50 (range <8 to 581.4). Whereas neutralizing titers predict the efficacy of the CCP in a transfusion recipient, the assays are impractical for the scale of the pandemic. In order to evaluate the ability of commercially available test kits to predict neutralizing capability, the total Ig to the S protein (Ortho-Vitros) and to the RBD (Lumit Dx, Promega) were measured. Whereas a majority of CCP units had low titers of anti-RBD antibodies, a few samples had higher anti-RBD titers. This trend was largely similar to the neutralizing titers of these samples. In agreement with the idea that anti-RBD antibodies are usually neutralizing (9), these assays were highly correlated (ρ of 0.8295) (Fig. 1A). Although the total anti S Ig as measured with the Ortho-Vitros assay also correlated reasonably with the IC50 values, the correlation coefficient was much lower (ρ =0.5637) (Fig. 1B). The Ortho-Vitros assay correlated with the anti-RBD antibody titers, as expected (ρ =0.6764) (Fig. 1C). These comparisons portray the value of the anti-RBD assay over other serological assays in identifying a CCP unit of higher neutralizing titer.

**Figure 1.**
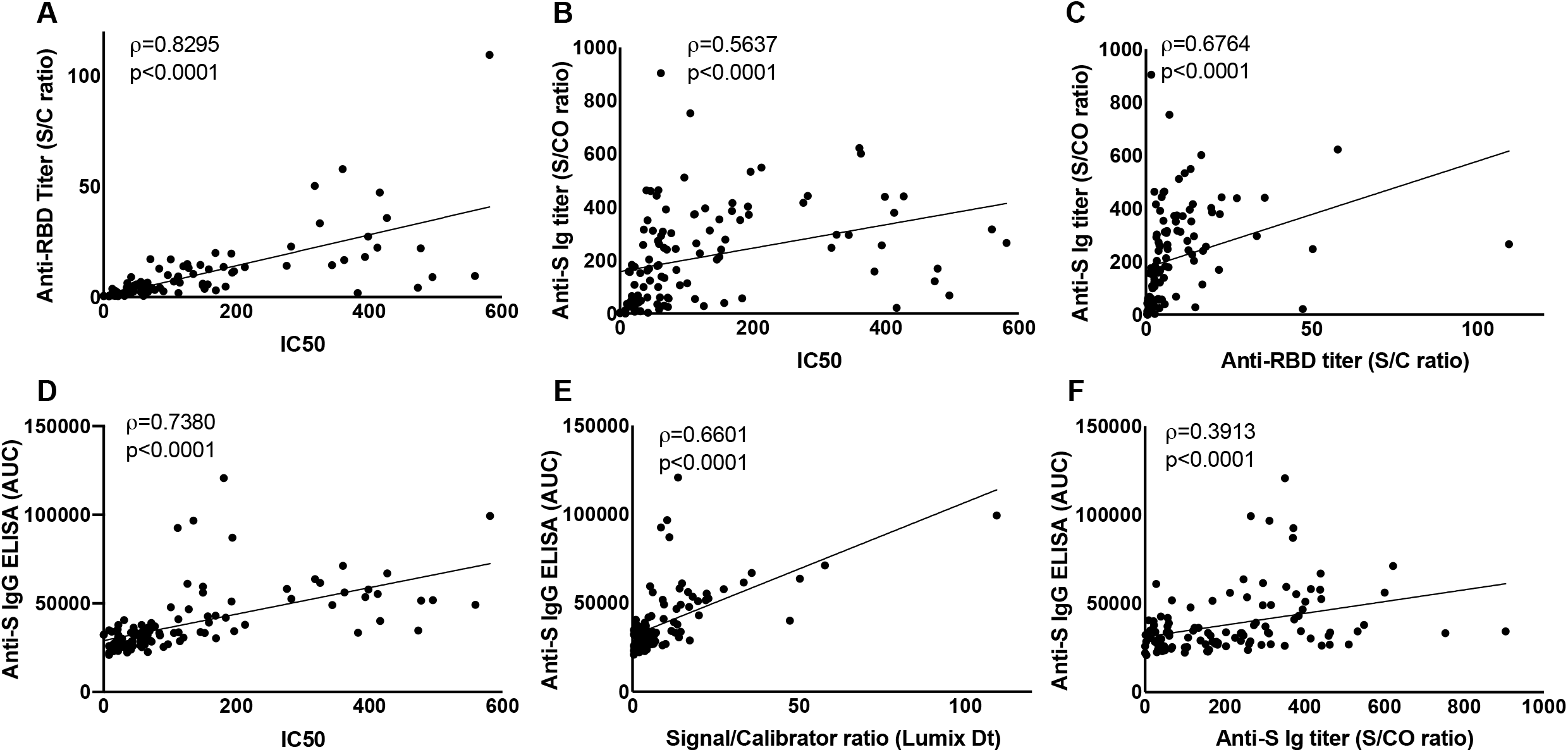
Correlative analysis of anti-SARS-CoV-2 immunoassays. Assays were performed to detect anti spike-Immunoglobulin (Ig) using the Ortho-Vitros assay (represented by signal to cut-off ratios (S/CO)), or anti-spike IgG with ELISA (represented by area under the curve, AUC), anti-RBD Ig using the Lumit Dx assay (represented by sample signal to calibrator ratios (S/C)) and 50% neutralizing titers with a retroviral-pseudotype assay (represented by IC50). Analysis of correlation were performed using Spearman’s correlation test for (A) anti-RBD titer, S/C vs IC50, (B) total anti-S Ig titer, S/CO vs IC50 and(C) anti-RBD titer, S/C vs total anti-S Ig titer, S/CO. These assay results were also qualified by comparing IC50 (D), anti-RBD titer, S/C (E) and total anti-S Ig titer, S/CO (F) against an in-house anti-spike protein IgG ELISA. Spearman’s correlation coefficient, ρ, and *p* values are indicated for each correlation.

Anti-S IgG, IgA and IgM were also measured with an ELISA developed in-house in the 111 CCP samples. The anti-S IgA and IgM did not correlate well with IC50 values or with the anti-RBD titers (data not shown). However, anti-S IgG strongly correlated with the IC50 titer (ρ =0.7380) (Fig. 1D) and with the anti-RBD titer (ρ =0.6601) (Fig. 1E). These correlations suggest that a majority of the neutralizing antibodies are likely to be of the IgG isotype. Perhaps the most surprising comparison showed that although, the anti-S IgG (Fig. 1F), IgA and IgM titers individually correlated with the total Ig titers from the Ortho-Vitros assay, these correlations were not as strong as those with the anti-RBD titers. This suggests that the Ortho-Vitros assay, which is designed to be qualitative, may not be amenable to accurate quantitation.

Taken together, these results confirm the idea that measuring anti-RBD antibodies is predictive of neutralizing titers to a greater degree than other assays. The therapeutic benefit of CCP may also be immediately evaluated by sampling transfusion recipients and measuring antibody titers before and after transfusion. The anti-RBD antibody titers in CCP recipients sampled longitudinally also confirmed the high degree of correlation between the two assays (ρ =0.8440) (Fig. 2A). Furthermore, by analyzing the anti-RBD titers in plasma recipients, transmissible antibody titers with the transfusion of CCP in at least 6 patients were observed (Fig. 2B).

**Figure 2.**
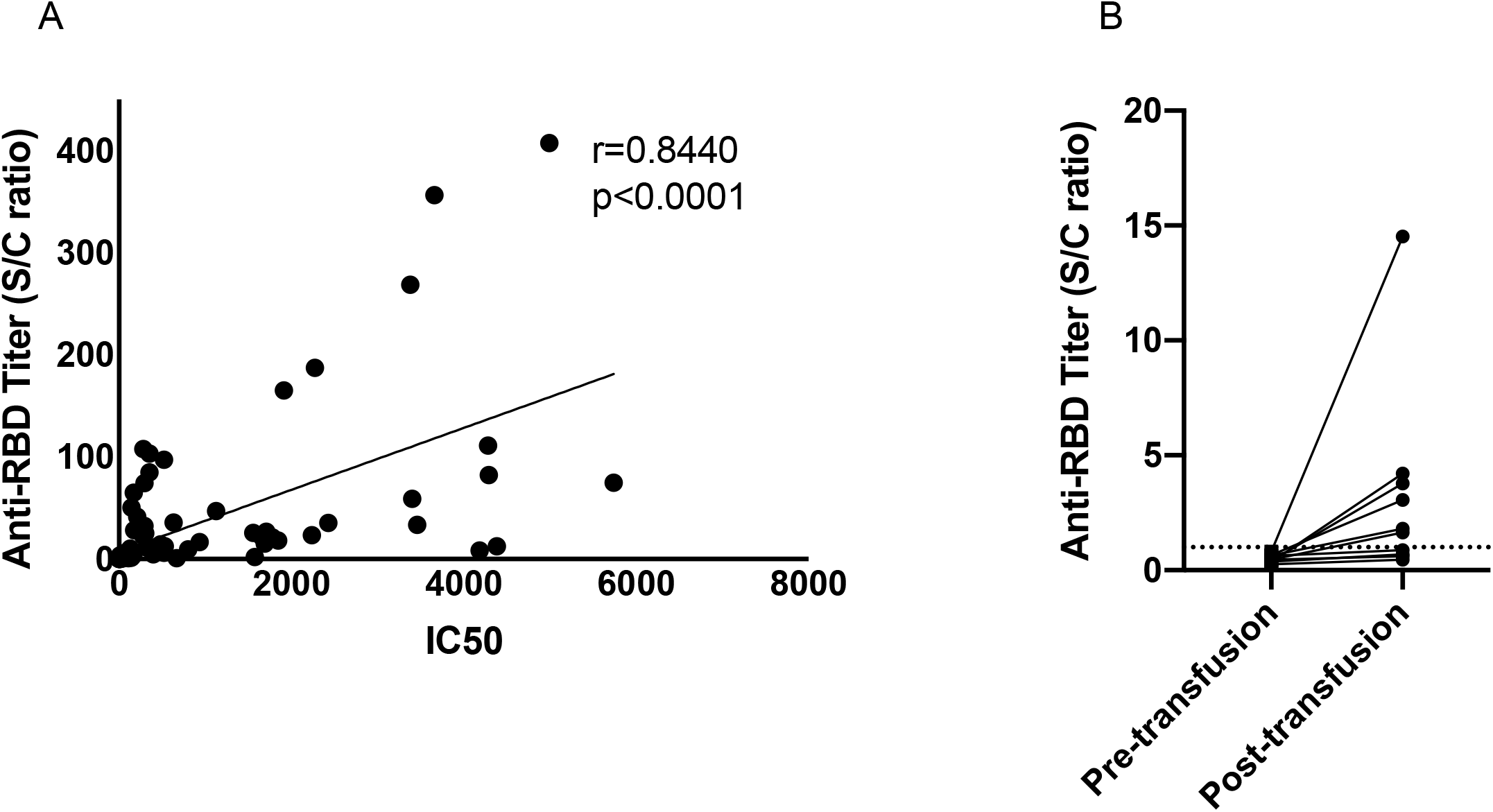
Neutralizing titer and anti-RBD antibody titers in CCP recipients, pre- and post-transfusion. (A) Plasma or serum samples from CCP recipients before or after transfusion were assayed for neutralizing ability and for anti-RBD Ig titers using the Lumit Dx assay. Correlation analysis was performed using Spearman’s test and the correlation coefficient, ρ, and *p* values are reported. (B) Plasma or serum samples from antibody naïve CCP recipients were analyzed for anti-RBD antibodies using the Lumit Dx assay system. Dashed line indicates the criteria to determine seropositivity of CCP units.

## Discussion

The antibody responses to SARS-CoV2 infection are highly variable in COVID-19 patients and several clinical characteristics have been associated with strong anti-SARS-CoV2 antibody production (11). Although these characteristics can be used to increase chances of selecting a CCP donor with the potential to produce a robust antibody response, the identification of a CCP with therapeutic value remains cumbersome. CCP with higher antibody titers that can neutralize the virus is essential to the success of plasma transfusion (4). Whereas analysis of neutralization is not practical in a clinical setting, several commercial assay platforms are currently available to test anti-SARS-CoV2 antibody titers. Of these, measures of anti-spike antibodies correlate best with the neutralizing titer (5, 7). In this study, using measures of anti-spike antibodies from three different platforms, and neutralizing capability of CCP, wide range of antibody responses in COVID-19 patients was confirmed.

Since virus neutralization is a necessary attribute for therapy (4), commercial assays were evaluated to test if they can predict the neutralizing ability of CCP. Several studies have documented the correlation of neutralization titers with different measures of antibodies in COVID-19 patients and with anti-RBD antibodies in particular (5-7, 12, 13). In studies where neutralizing titers of antibodies were not known, or were predicted with sub-optimal assay platforms, the effect of high antibody titer CCP has likely been underestimated (14, 15). In this study, the Lumit Dx assay system was evaluated as a surrogate system for detecting anti-RBD antibodies and to predict CCP units with higher neutralization titers.

The presence of neutralizing antibodies can be detected in patients as early as one week after the onset of symptoms (16). In approximately 1% of patients, where the neutralizing titer is extremely high within 7 days post onset of symptoms, neutralizing antibodies are detectable at high levels at least 2 months later (17). On the other hand, in patients with lower titers early on, neutralizing activity diminishes soon after (17). These trends are largely in line with antibody measurements against the S or the nucleocapsid protein of SARS-CoV2 (7, 13, 18, 19). The duration of the neutralization response is an important consideration when evaluating plasma for transfusion therapy (20). Transfusion of CCP with lower antiviral activity could result in the generation of neutralization escape variants, and also affect the likelihood of re-infection in plasma recipients. In several studies, antibody levels required to theoretically expect a positive intervention were not met (4). In a previous study, we have reported that up to 50% of locally collected CCP units may have the neutralizing activity necessary (8) and this study extends these observations.

Other systems to measure anti-RBD are also available commercially (13). The importance of these assay platforms lies in the global scale of the pandemic and the necessity to quickly evaluate and deploy convalescent plasma for treatment. While the efficacy of the different vaccines against newer and more contagious strains of SARS-CoV2 are evaluated, it is necessary to maximize the use of available treatment modes. To effectively make decisions to manage the scale of the pandemic, an assay platform that is quick, and deployable with limited training is crucial. Of note, the Lumit Dx assay can provide a measure of anti-RBD antibodies, and neutralizing potential, in 45 minutes and can be scaled for a high throughput format of the assay. This assay can also be automated with a liquid-handling robot to further increase throughput.

In conclusion, with the analysis of over 200 clinical samples, we demonstrate the utility of the Lumit Dx assay in predicting the neutralizing ability of CCP. The convenience of the Lumit Dx assay system makes the measurement of anti-RBD antibodies amenable to a high throughput setup with a rapid turnaround time. Despite the assay-design being qualitative, the dynamic range of the output extends over several decades and thus can be used to extrapolate a quantitative readout of anti-RBD antibody titers.

## Data Availability

Can be made available after contact with authors

## Acknowledgment

This study was funded by Research and Development funds provided to Joseph Connor, MD by the University of Wisconsin Department of Pathology and Laboratory Medicine.

The authors would like to thank the University of Wisconsin Carbone Cancer Center (UWCCC) for use of its Shared Services to complete this research. This work is supported in part by NIH/NCI P30 CA014520-UW Comprehensive Cancer Center Support. The ability of the BioBank to mobilize these COVID-19 research effort is based on leveraging established relationships with outstanding colleagues and departments including the Department of Pathology & Laboratory Medicine, the Institutional Review Board (IRB), the Institute of Clinical and Translational Research (ICTR) and the UW school of Medicine and Public Health (SMPH). Lumit Dx assay reagents were provided by Promega Corporation (Madison, WI). No members of the Promega Organization were involved in the study design, experiments and interpretation of data.

Lastly, the authors would like to thank Drs. Susan Stramer and Erin Goodhue of the American Red Cross for permitting the use of samples from a previous collaborative study. The use of these samples for the current study was reviewed and approved by the IRB of the ARC.

